## Supplementary material for "Unbiased single cell spatial analysis localises inflammatory clusters of immature neutrophils-CD8 T cells to alveolar progenitor cells in fatal COVID-19 lungs": All Suppl figures

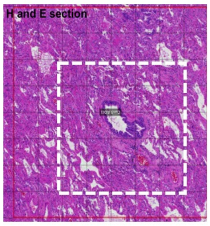


A. Analysis of overall histo-pathology features from HnE stained first slide from paraffin embedded tissue. Relevant ROI drawn (2-3 of 2x2mm per patient).

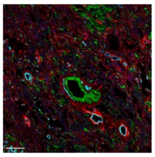

B. Staining of adjacent lung tissue section with a 37-plex metal-tagged antibody panel and acquisition by IMC (Hyperion™). Individual metal intensity signals linked to x,y coordinate of ablated area are then digitised to generate an MCD image file.

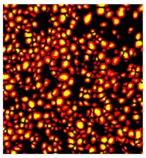

C. Segmentation of OME-TIFF (generated from MCD image file) into single cells. Cell masks and metadata (x,y location, area) are generated using Mesmer library from Deep Cell platform (Greenwald N 2022)

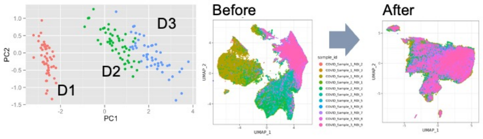


D. Signal intensity from each cell is extracted for dimension reduction and clustering using R Phenograph. Batch effect from ablation and staining days is adjusted using Harmony integration (Korsunsky I 2021).

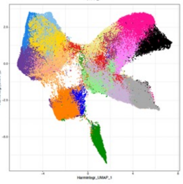

E. Annotation of clusters is performed using expression density plot, heat map analysis, Pseudotime of selected groups of clusters, cluster distribution plot, and cross checked with H&E and MCD images (Figure S7).

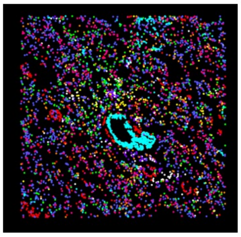

F. Creation of cell centroid map, providing identity and location for all annotated cells in a given ROI.

### Spatial statistical analysis of co-locating cells

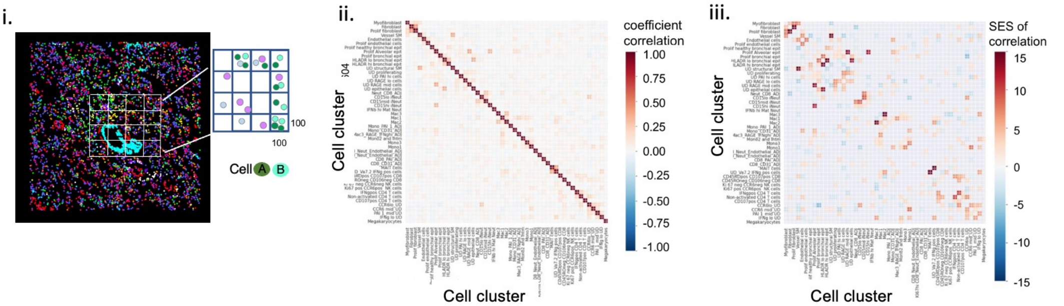


**G. Step 1. Quadrat Correlation Matrix (QCM) analysis.** Each ROI is first divided into 100x100um quadrats (i, only 16 quadrats out of 400 quadrats in the 2x2 mm ROI are shown, for clarity) and correlation between all pairs of annotated cells in each quadrat is analyzed using Morueta-Holme 2016 methods. Coefficients of correlation and standardized effect size (SES) for these correlations (above null distribution) and adjusted for multiple comparisons by Bonferroni correction are calculated (ii and iii). Pairs of cells that have SES with FDR q < 0.05 are submitted for cross pair correlation function (cross-PCF) analysis.

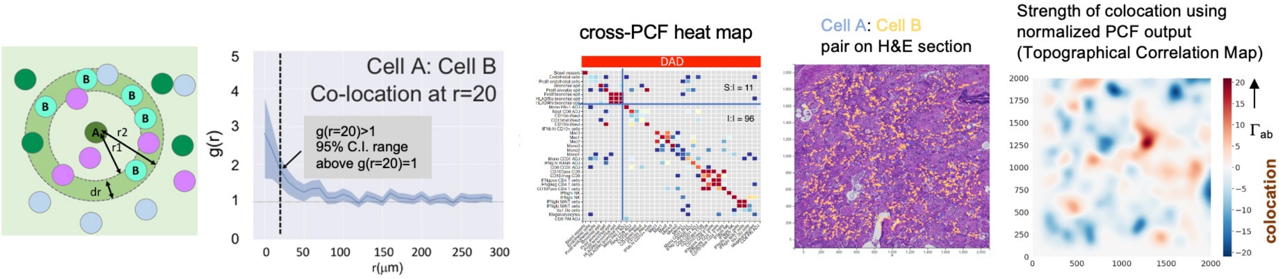


**H. Step 2. Cross pair correlation function (cross-PCF) analysis.** Pairs of cells which passed the significant correlation test from the QCM analysis are tested for co-location with each other. An annulus is drawn round an anchor cell of interest (cell A) at r1=20μm and r2 =30μm from the centroid of cell A. Frequency of cell B in this annulus is calculated against complete spatial randomness and a 95% confidence interval fitted for the curve for pairs of cells in a specified relevant group of ROIs. Pairs of cells with g(r=20) >1 and 95% C.I. range above g(r=20)=1 are termed statistically significantly co-located at 20μm from the designated anchor cell (Cell A). Finally, a topographical representation of statistically significant co-locating cells is generated for a given ROI using an adapted PCF, termed '$\Gamma_{ab}$'. $\Gamma_{ab}$ measures the frequency of cells in the space from the centroid of Cell A to the edge of r2.

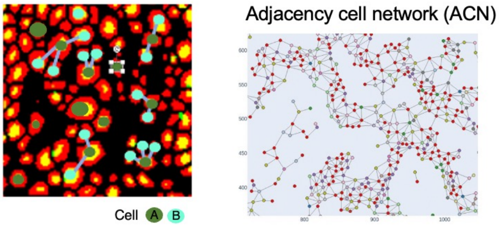


**I. Step 3. Adjacent cell network (ACN).** For the cell pairs that were significantly co-located, we then analyze the network of cells that are in direct physical contact with the anchor cell of interest using a boundary enhanced cell centroid map. Thus, for cell A (anchor cell) that has been found to be significantly co-located with cell B by QCM and cross-PCF analysis, we compute the proportion of cell A in the ROI that are physically in contact with cell B ($\phi_{AB}$ , see methods). In this exemplar plot, there are 12 cells of type A in the ROI; and 8 (67%) have contact with at least one cell of type B.

Figure S1

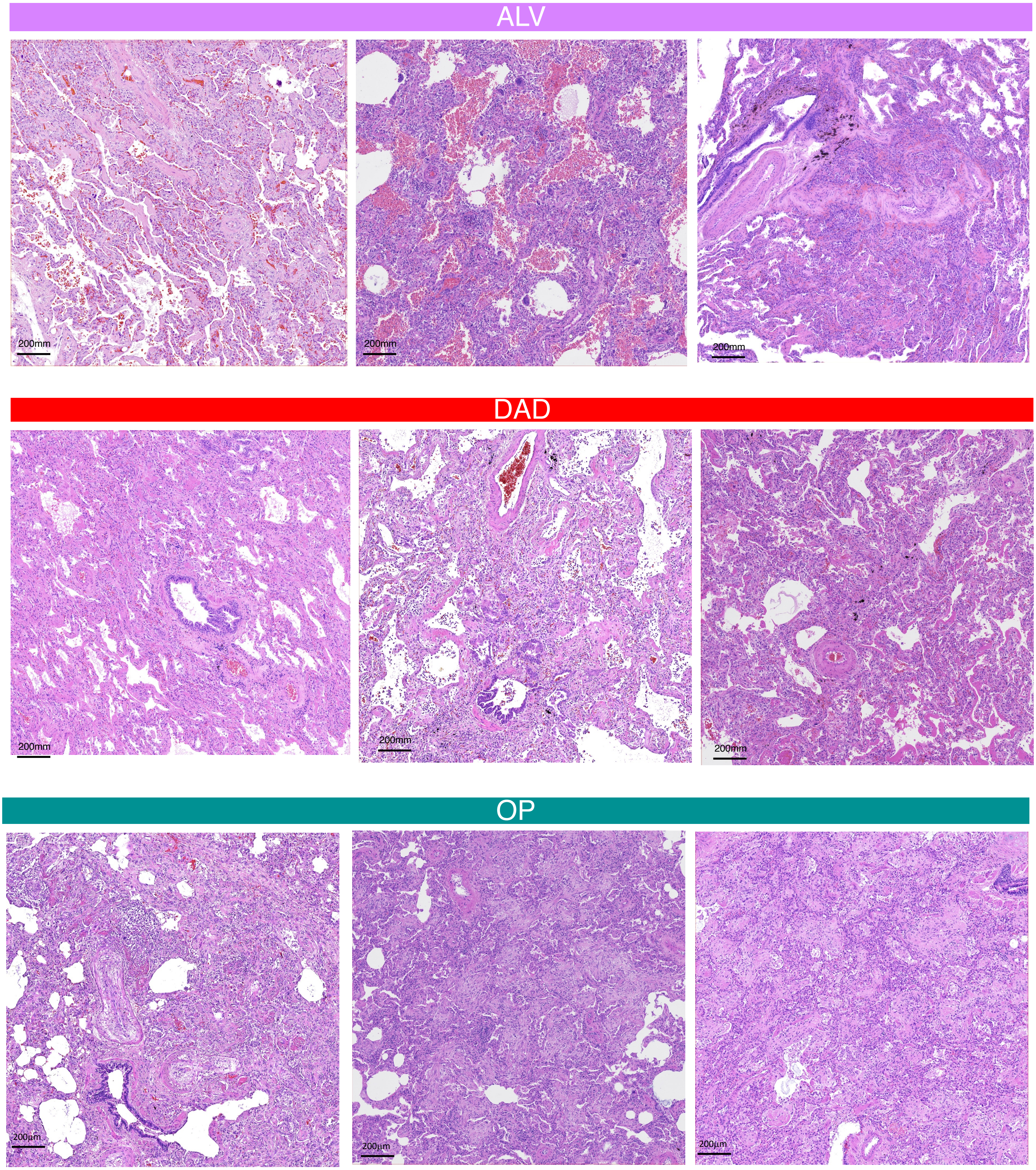


Figure S2

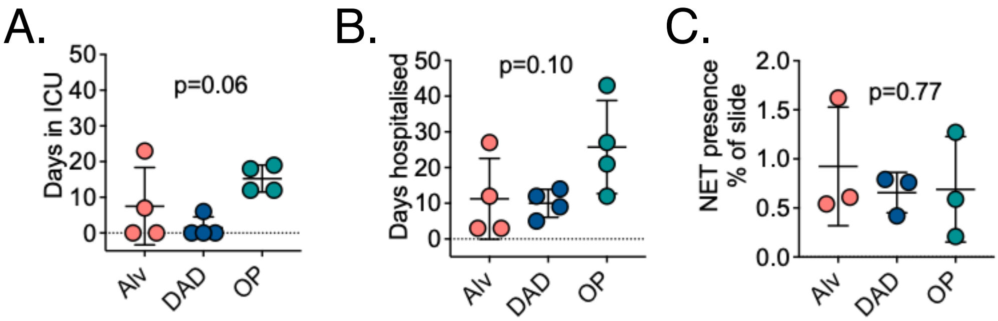
A.
Days in ICU
p=0.06
B.
Days hospitalised
p=0.10
C.
NET presence
% of slide
p=0.77
Alv
DAD
OP

Figure S3

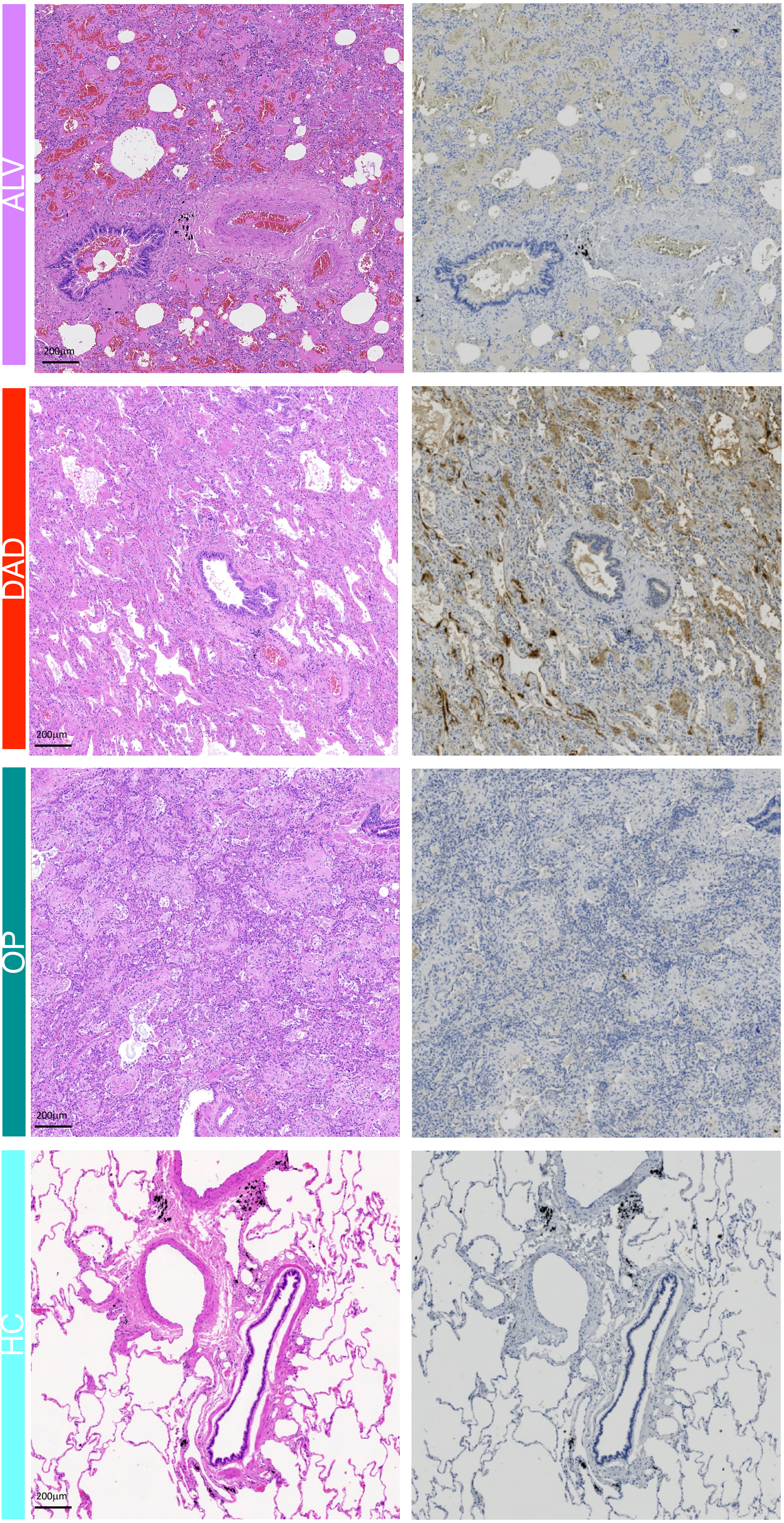


Figure S4

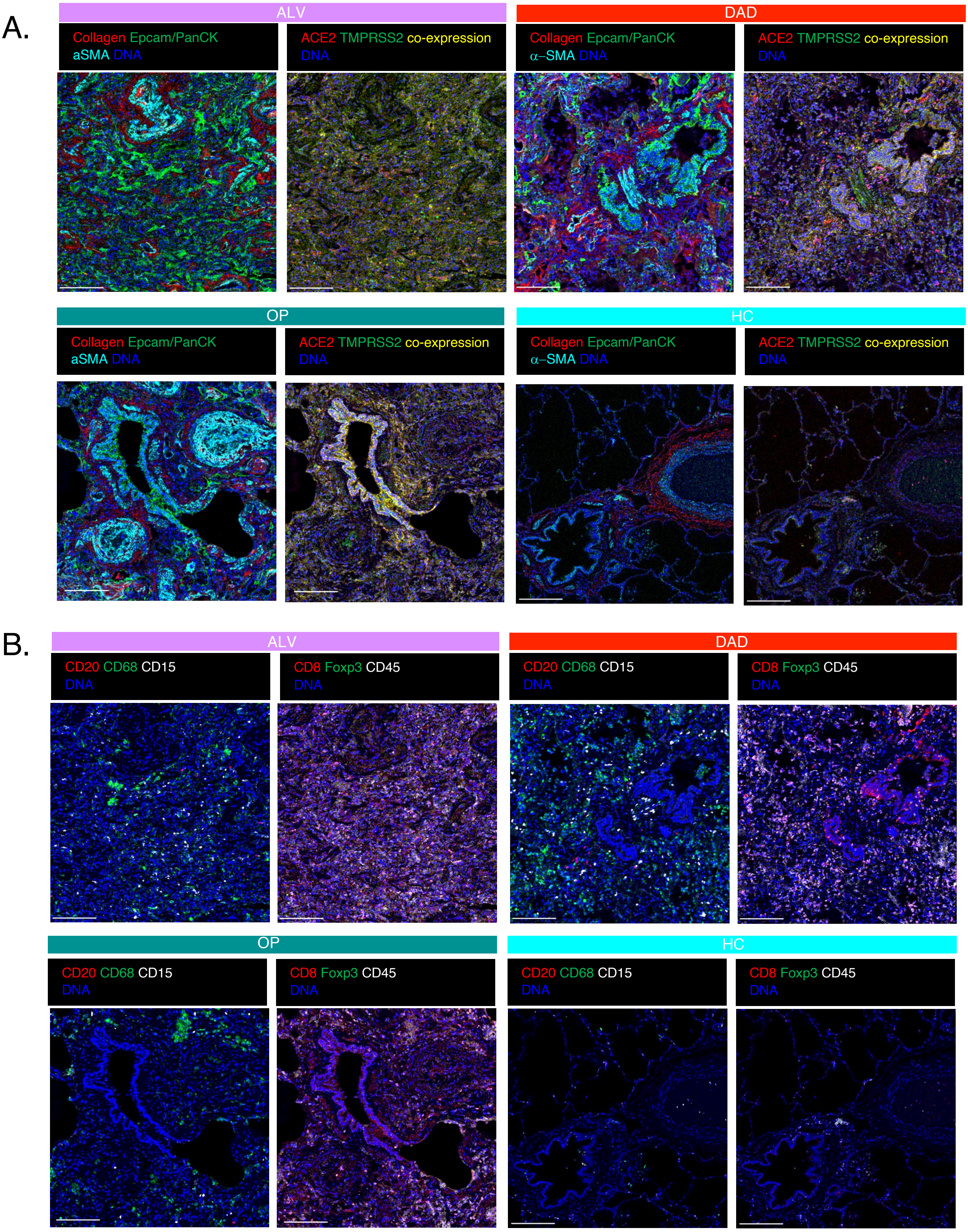

A.
ALV
DAD
Collagen Epcam/PanCK
aSMA DNA
ACE2 TMPRSS2 co-expression
DNA
Collagen Epcam/PanCK
α–SMA DNA
ACE2 TMPRSS2 co-expression
DNA
OP
HC
Collagen Epcam/PanCK
aSMA DNA
ACE2 TMPRSS2 co-expression
DNA
Collagen Epcam/PanCK
α–SMA DNA
ACE2 TMPRSS2 co-expression
DNA
B.
ALV
DAD
CD20 CD68 CD15
DNA
CD8 Foxp3 CD45
DNA
CD20 CD68 CD15
DNA
CD8 Foxp3 CD45
DNA
OP
HC
CD20 CD68 CD15
DNA
CD8 Foxp3 CD45
DNA
CD20 CD68 CD15
DNA
CD8 Foxp3 CD45
DNA


Figure S5

A.

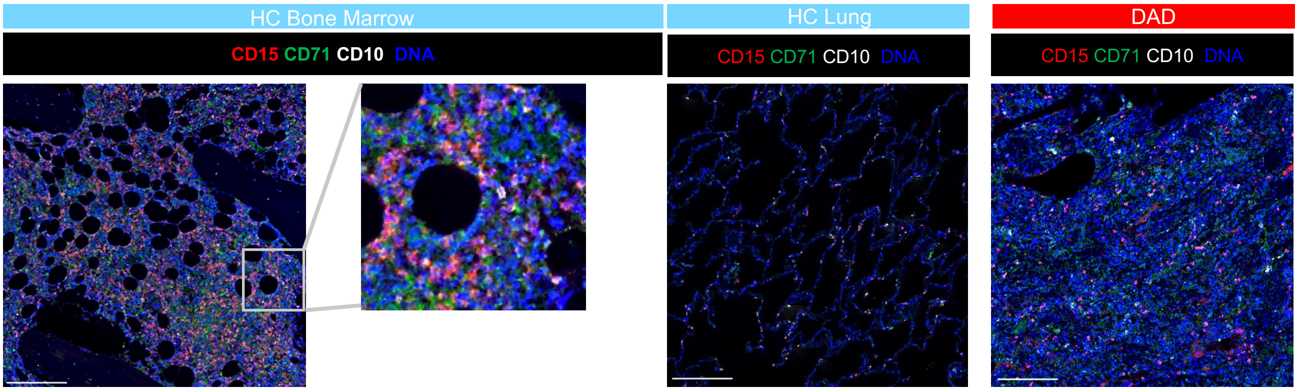


B.

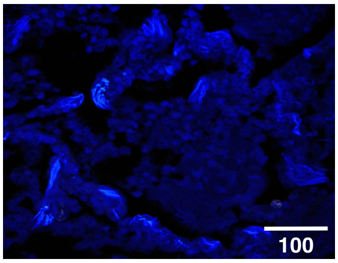


C.

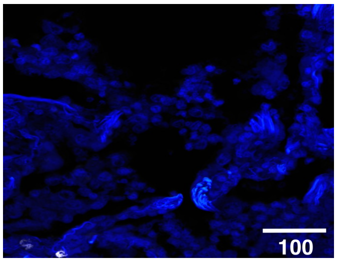


Figure S6

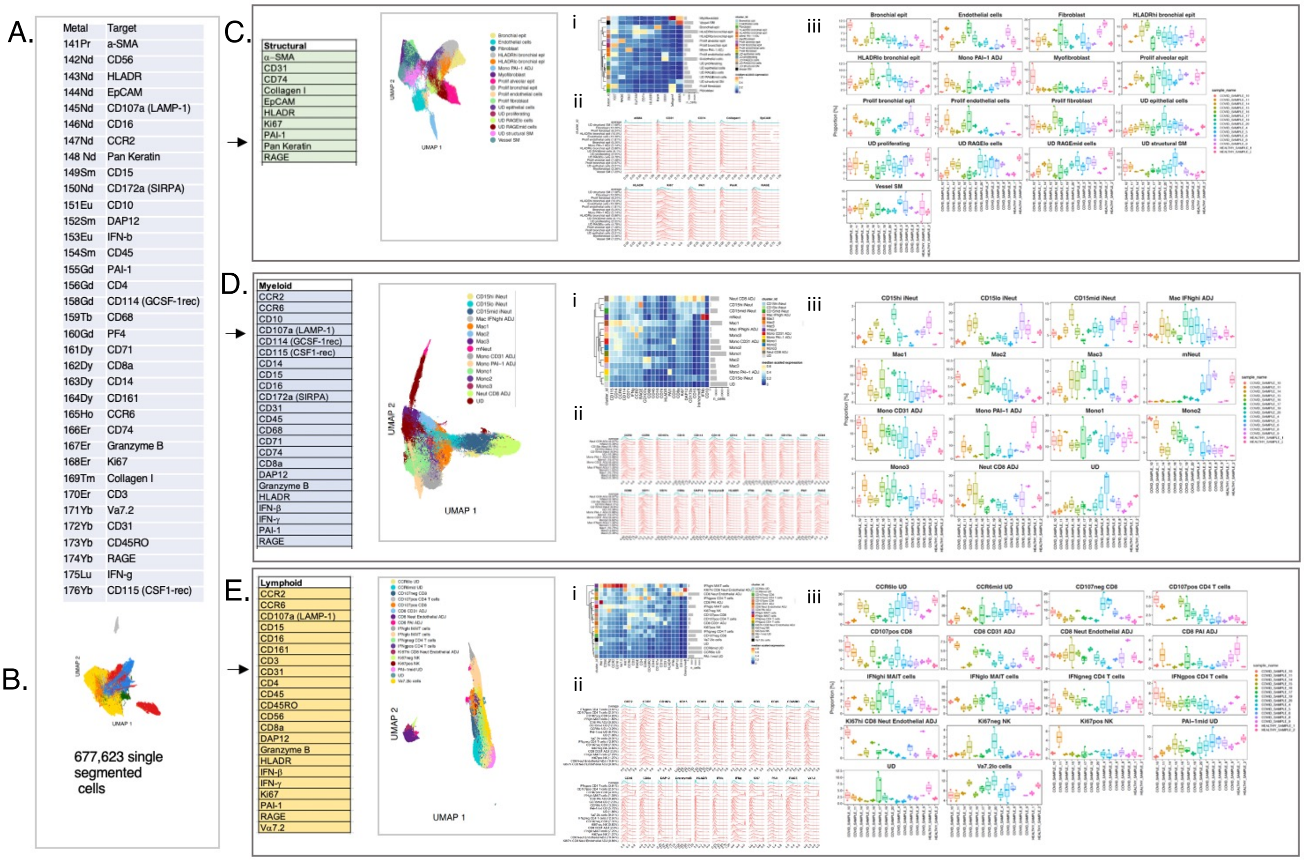
A.
Metal Target
141Pr a-SMA
142Nd CD56
143Nd HLADR
144Nd EpCAM
145Nd CD107a (LAMP-1)
146Nd CD16
147Nd CCR2
148 Nd Pan Keratin
149Sm CD15
150Nd CD172a (SIRPA)
151Eu CD10
152Sm DAP12
153Eu IFN-b
154Sm CD45
155Gd PAI-1
156Gd CD4
158Gd CD114 (GCSF-1rec)
159Tb CD68
160Gd PF4
161Dy CD71
162Dy CD8a
163Dy CD14
164Dy CD161
165Ho CCR6
166Er CD74
167Er Granzyme B
168Er Ki67
169Tm Collagen I
170Er CD3
171Yb Va7.2
172Yb CD31
173Yb CD45RO
174Yb RAGE
175Lu IFN-g
176Yb CD115 (CSF1-rec)
B.
677,623 single segmented cells
C.
Structural
α-SMA
CD31
CD74
Collagen I
EpCAM
HLADR
Ki67
PAI-1
Pan Keratin
RAGE
UMAP 1
UMAP 2
i
ii
iii
D.
Myeloid
CCR2
CCR6
CD10
CD107a (LAMP-1)
CD114 (GCSF-1rec)
CD115 (CSF1-rec)
CD14
CD15
CD16
CD172a (SIRPA)
CD31
CD45
CD68
CD71
CD74
CD8a
DAP12
Granzyme B
HLADR
IFN-β
IFN-γ
PAI-1
RAGE
UMAP 1
UMAP 2
i
ii
iii
E.
Lymphoid
CCR2
CCR6
CD107a (LAMP-1)
CD15
CD16
CD161
CD3
CD31
CD4
CD45
CD45RO
CD56
CD8a
DAP12
Granzyme B
HLADR
IFN-β
IFN-γ
Ki67
PAI-1
RAGE
Vα7.2
UMAP 1
UMAP 2
i
ii
iii

Figure S7

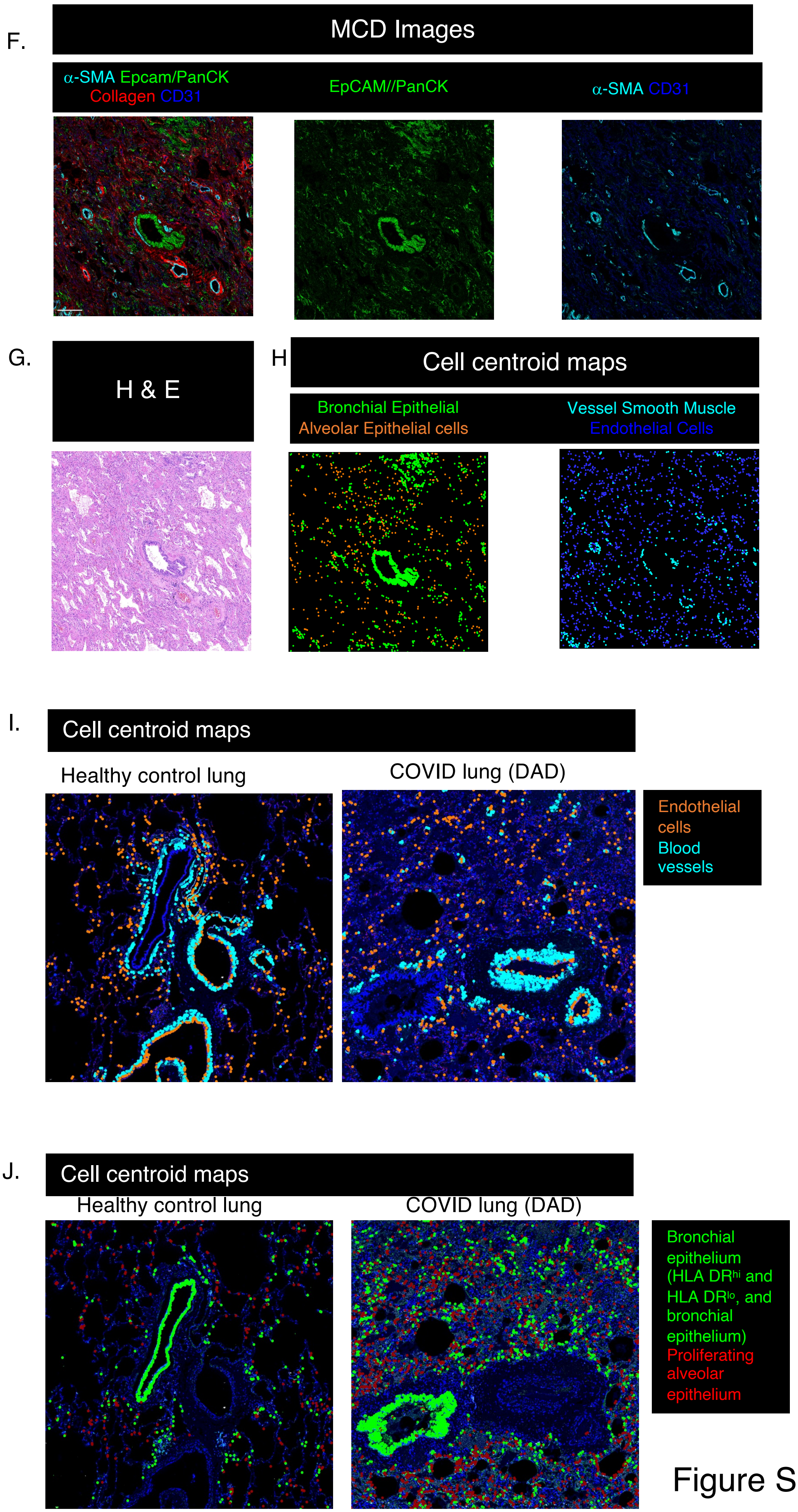
F.
MCD Images
α-SMA Epcam/PanCK
Collagen CD31
EpCAM//PanCK
α-SMA CD31
G.
H & E
H
Cell centroid maps
Bronchial Epithelial
Alveolar Epithelial cells
Vessel Smooth Muscle
Endothelial Cells
I.
Cell centroid maps
Healthy control lung
COVID lung (DAD)
Endothelial cells
Blood vessels
J.
Cell centroid maps
Healthy control lung
COVID lung (DAD)
Bronchial epithelium (HLA DR$^{hi}$ and HLA DR$^{lo}$, and bronchial epithelium)
Proliferating alveolar epithelium

Figure S7

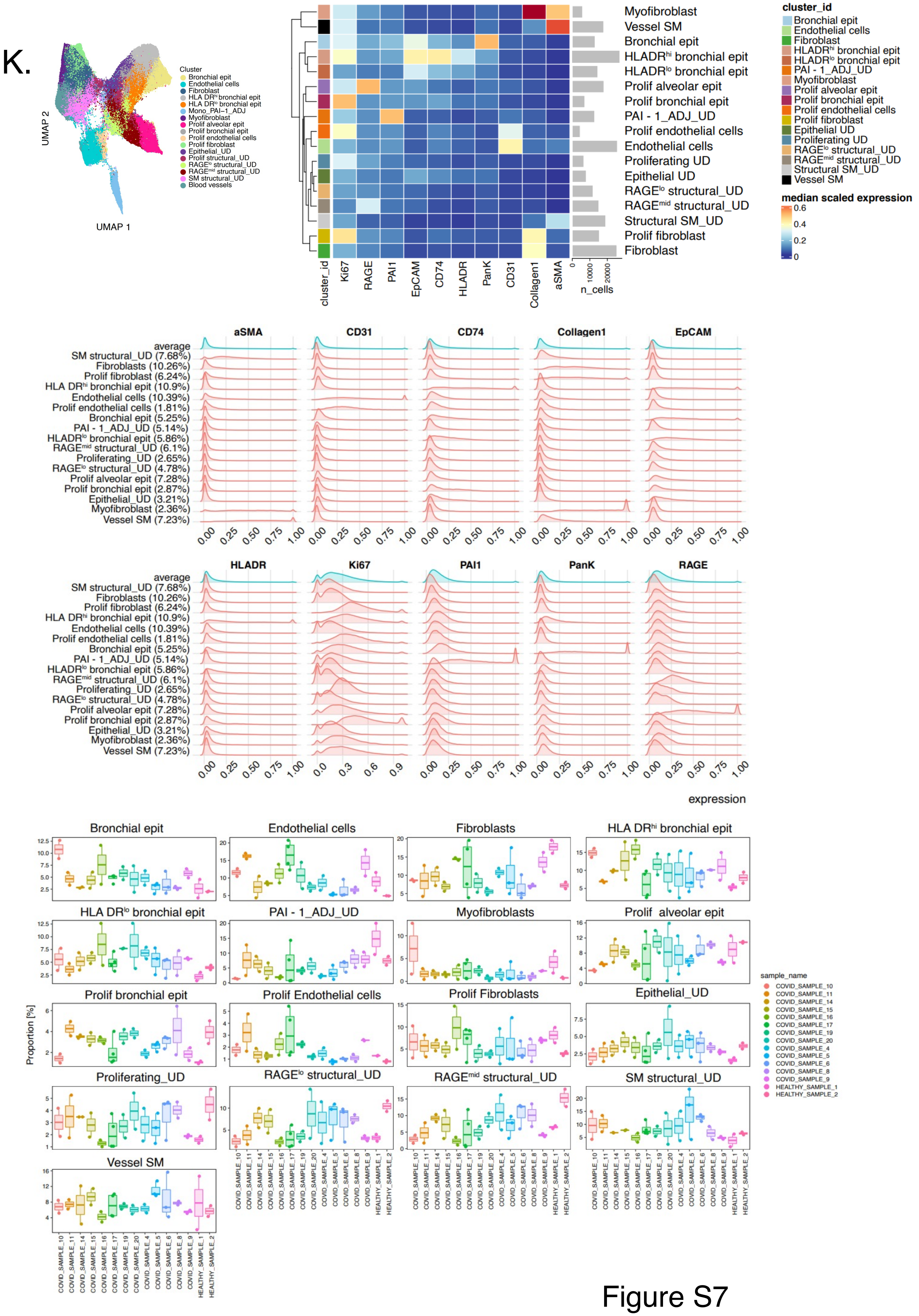
K.
UMAP 2
UMAP 1
Cluster
Bronchial epit
Endothelial cells
Fibroblast
HLA DR$^{hi}$ bronchial epit
HLA DR$^{lo}$ bronchial epit
Mono_PAI−1_ADJ
Myofibroblast
Prolif alveolar epit
Prolif bronchial epit
Prolif endothelial cells
Prolif fibroblast
Epithelial_UD
Prolif structural_UD
RAGE$^{lo}$ structural_UD
RAGE$^{mid}$ structural_UD
SM structural_UD
Blood vessels
Myofibroblast
Vessel SM
Bronchial epit
HLADR$^{hi}$ bronchial epit
HLADR$^{lo}$ bronchial epit
Prolif alveolar epit
Prolif bronchial epit
PAI - 1_ADJ_UD
Prolif endothelial cells
Endothelial cells
Proliferating UD
Epithelial UD
RAGE$^{lo}$ structural_UD
RAGE$^{mid}$ structural_UD
Structural SM_UD
Prolif fibroblast
Fibroblast
cluster_id
Ki67
RAGE
PAI1
EpCAM
CD74
HLADR
PanK
CD31
Collagen1
aSMA
n_cells
median scaled expression
aSMA
CD31
CD74
Collagen1
EpCAM
HLADR
Ki67
PAI1
PanK
RAGE
average
SM structural_UD (7.68%)
Fibroblasts (10.26%)
Prolif fibroblast (6.24%)
HLA DR$^{hi}$ bronchial epit (10.9%)
Endothelial cells (10.39%)
Prolif endothelial cells (1.81%)
Bronchial epit (5.25%)
PAI - 1_ADJ_UD (5.14%)
HLADR$^{lo}$ bronchial epit (5.86%)
RAGE$^{mid}$ structural_UD (6.1%)
Proliferating_UD (2.65%)
RAGE$^{lo}$ structural_UD (4.78%)
Prolif alveolar epit (7.28%)
Prolif bronchial epit (2.87%)
Epithelial_UD (3.21%)
Myofibroblast (2.36%)
Vessel SM (7.23%)
expression
Bronchial epit
Endothelial cells
Fibroblasts
HLA DR$^{hi}$ bronchial epit
HLA DR$^{lo}$ bronchial epit
PAI - 1_ADJ_UD
Myofibroblasts
Prolif alveolar epit
Prolif bronchial epit
Prolif Endothelial cells
Prolif Fibroblasts
Epithelial_UD
Proliferating_UD
RAGE$^{lo}$ structural_UD
RAGE$^{mid}$ structural_UD
SM structural_UD
Vessel SM
Proportion [%]
sample_name
COVID_SAMPLE_10
COVID_SAMPLE_11
COVID_SAMPLE_14
COVID_SAMPLE_15
COVID_SAMPLE_16
COVID_SAMPLE_17
COVID_SAMPLE_19
COVID_SAMPLE_20
COVID_SAMPLE_4
COVID_SAMPLE_5
COVID_SAMPLE_6
COVID_SAMPLE_8
COVID_SAMPLE_9
HEALTHY_SAMPLE_1
HEALTHY_SAMPLE_2

Figure S7

L.

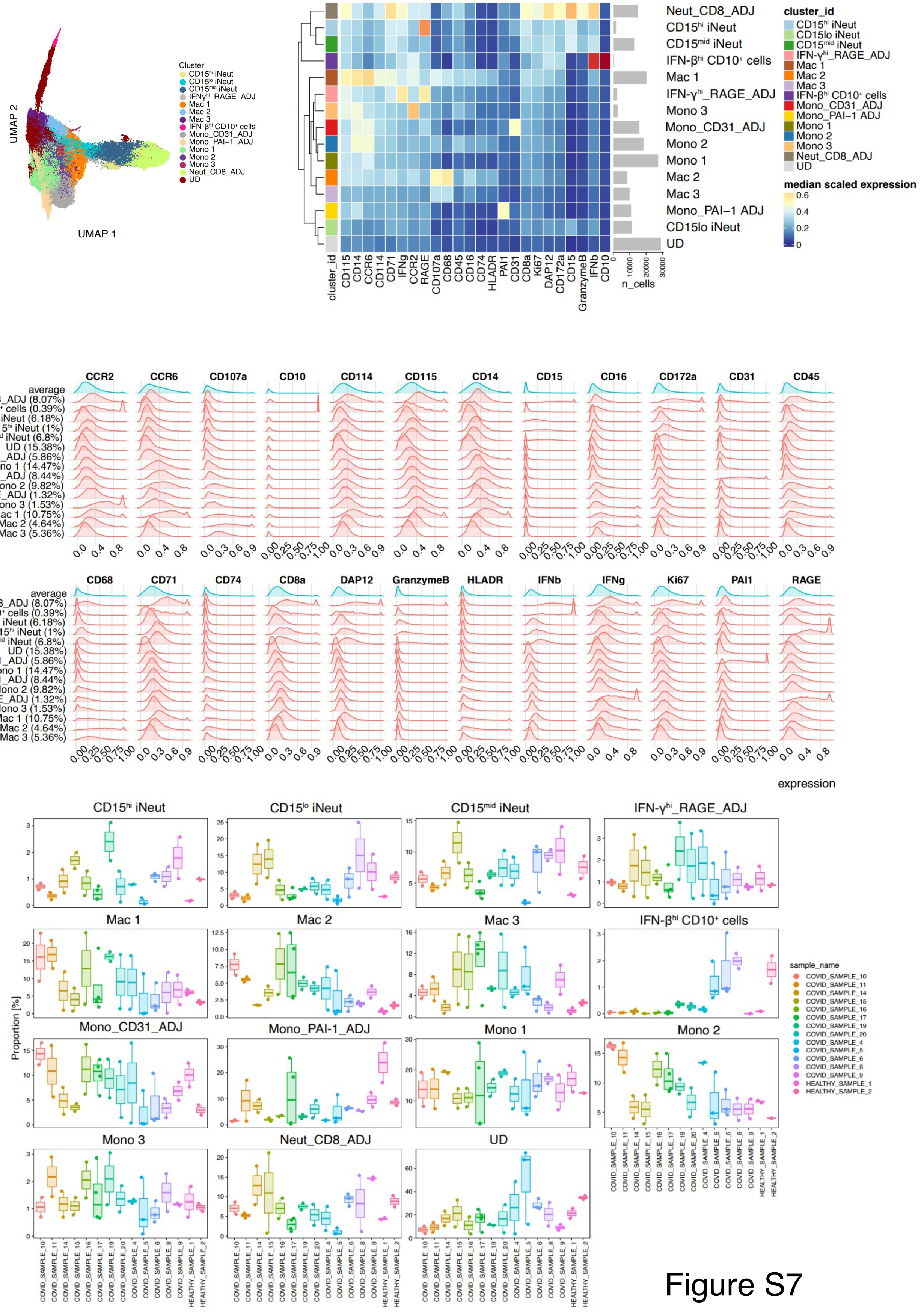

UMAP 2
UMAP 1
Cluster
CD15hi iNeut
CD15lo iNeut
CD15mid iNeut
IFNγhi_RAGE_ADJ
Mac 1
Mac 2
Mac 3
IFN-βhi CD10+ cells
Mono_CD31_ADJ
Mono_PAI-1_ADJ
Mono 1
Mono 2
Mono 3
Neut_CD8_ADJ
UD
cluster_id
n_cells
median scaled expression
CD115 CD14 CCR6 CD114 CD71 IFNg CCR2 RAGE CD107a CD68 CD45 CD16 CD74 HLADR PAI1 CD31 CD8a Ki67 DAP12 CD172a CD15 GranzymeB IFNb CD10
average
Neut_CD8_ADJ (8.07%)
IFN-βhi CD10+ cells (0.39%)
CD15lo iNeut (6.18%)
CD15hi iNeut (1%)
CD15mid iNeut (6.8%)
UD (15.38%)
Mono_PAI-1_ADJ (5.86%)
Mono 1 (14.47%)
Mono_CD31_ADJ (8.44%)
Mono 2 (9.82%)
IFN-γhi_RAGE_ADJ (1.32%)
Mono 3 (1.53%)
Mac 1 (10.75%)
Mac 2 (4.64%)
Mac 3 (5.36%)
expression
Proportion [%]
sample_name
COVID_SAMPLE_10
COVID_SAMPLE_11
COVID_SAMPLE_14
COVID_SAMPLE_15
COVID_SAMPLE_16
COVID_SAMPLE_17
COVID_SAMPLE_19
COVID_SAMPLE_20
COVID_SAMPLE_4
COVID_SAMPLE_5
COVID_SAMPLE_6
COVID_SAMPLE_8
COVID_SAMPLE_9
HEALTHY_SAMPLE_1
HEALTHY_SAMPLE_2


Figure S7

M.

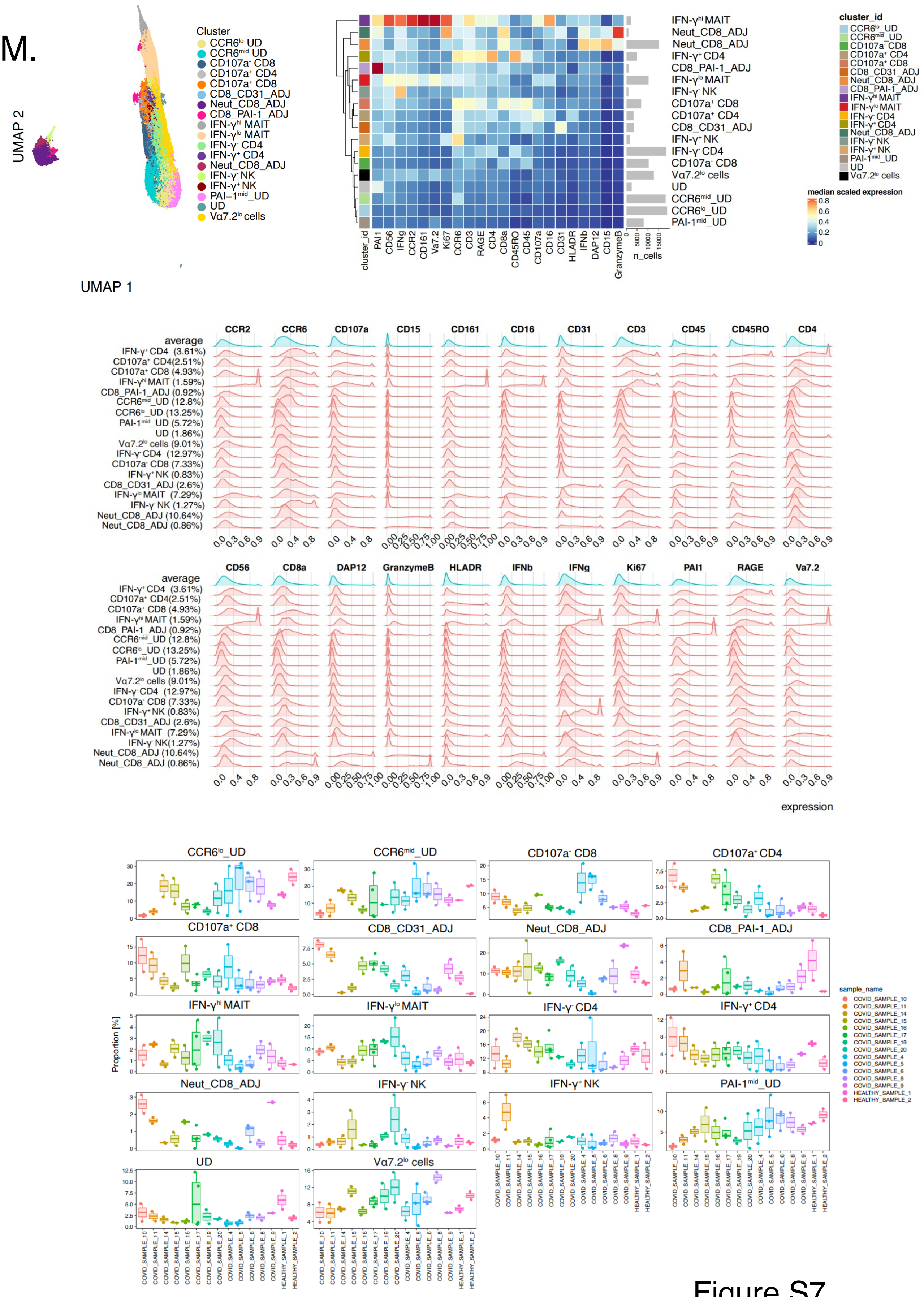
UMAP 2
UMAP 1
Cluster
CCR6lo UD
CCR6mid UD
CD107a- CD8
CD107a+ CD4
CD107a+ CD8
CD8_CD31_ADJ
Neut_CD8_ADJ
CD8_PAI-1_ADJ
IFN-γhi MAIT
IFN-γlo MAIT
IFN-γ- CD4
IFN-γ+ CD4
Neut_CD8_ADJ
IFN-γ- NK
IFN-γ+ NK
PAI-1mid_UD
UD
Vα7.2lo cells
cluster_id
n_cells
median scaled expression
average
expression
Proportion [%]
sample_name
COVID_SAMPLE_10
COVID_SAMPLE_11
COVID_SAMPLE_14
COVID_SAMPLE_15
COVID_SAMPLE_16
COVID_SAMPLE_17
COVID_SAMPLE_19
COVID_SAMPLE_20
COVID_SAMPLE_4
COVID_SAMPLE_5
COVID_SAMPLE_6
COVID_SAMPLE_8
COVID_SAMPLE_9
HEALTHY_SAMPLE_1
HEALTHY_SAMPLE_2

Figure S7

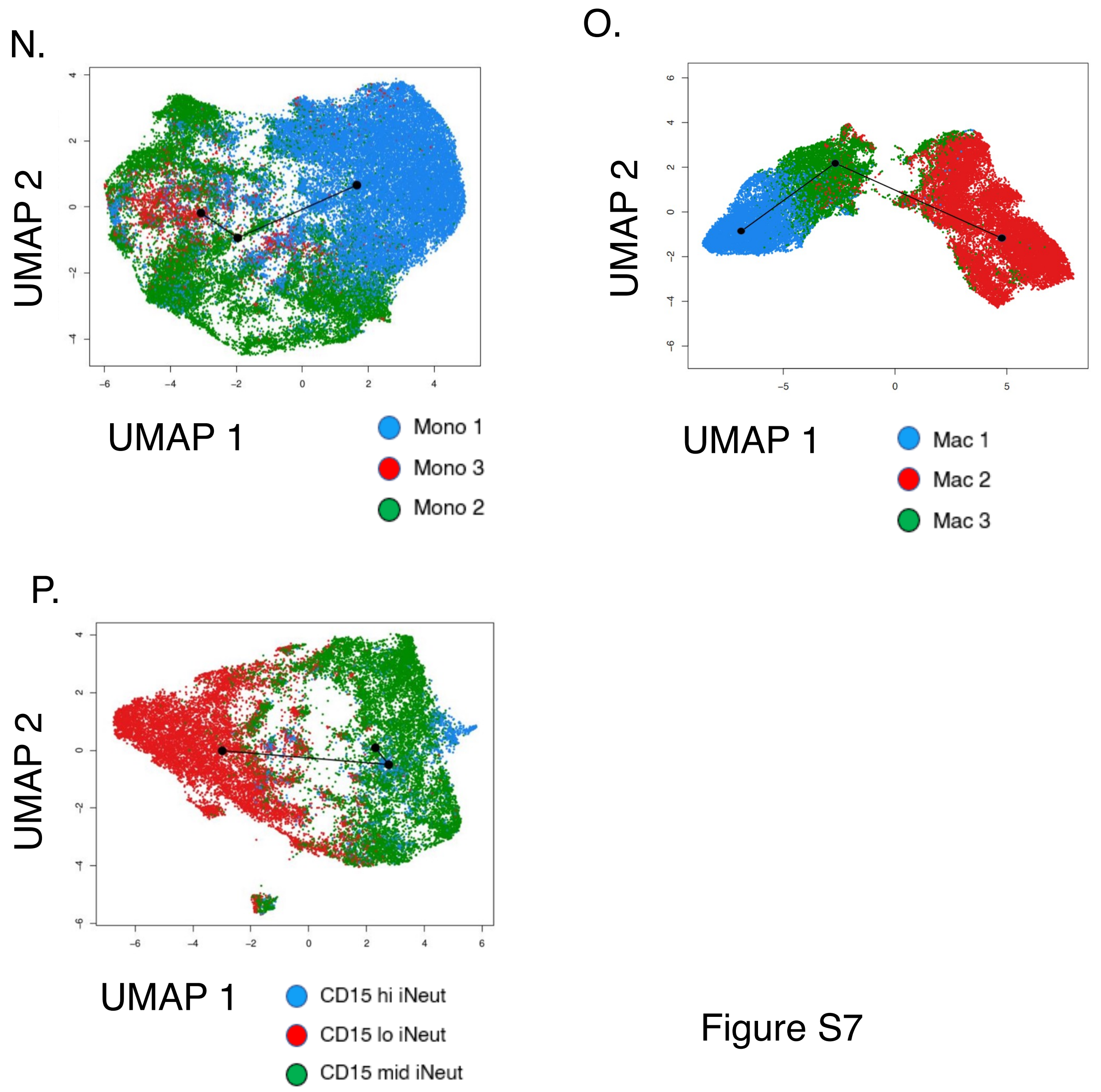
N.
UMAP 2
UMAP 1
Mono 1
Mono 3
Mono 2
O.
UMAP 2
UMAP 1
Mac 1
Mac 2
Mac 3
P.
UMAP 2
UMAP 1
CD15 hi iNeut
CD15 lo iNeut
CD15 mid iNeut

Figure S7

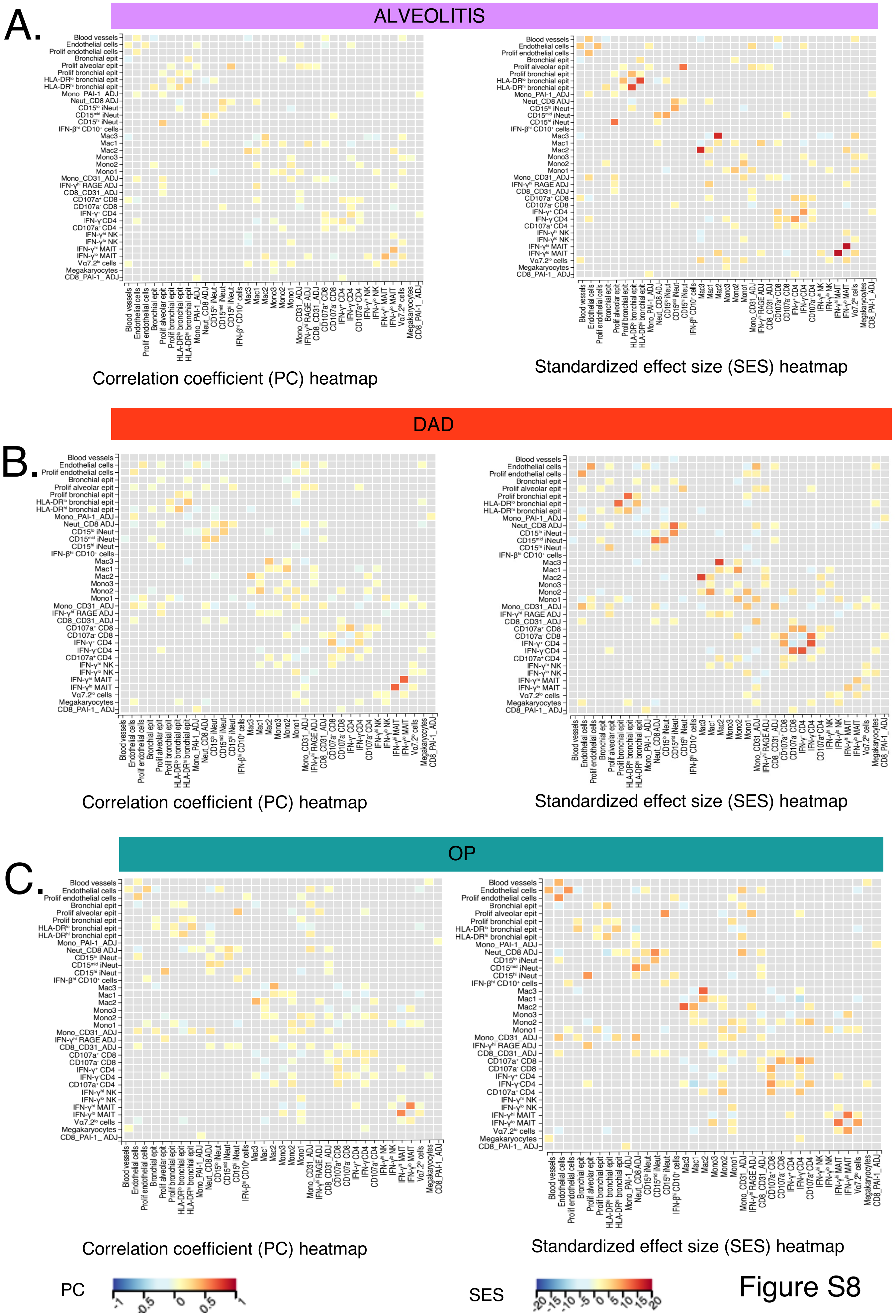
A.
ALVEOLITIS
Blood vessels
Endothelial cells
Prolif endothelial cells
Bronchial epit
Prolif alveolar epit
Prolif bronchial epit
HLA-DR^lo bronchial epit
HLA-DR^hi bronchial epit
Mono_PAI-1_ADJ
Neut_CD8 ADJ
CD15^lo iNeut
CD15^mid iNeut
CD15^hi iNeut
IFN-β^hi CD10+ cells
Mac3
Mac1
Mac2
Mono3
Mono2
Mono1
Mono_CD31_ADJ
IFN-γ^hi RAGE ADJ
CD8_CD31_ADJ
CD107a+ CD8
CD107a- CD8
IFN-γ+ CD4
IFN-γ-CD4
CD107a+ CD4
IFN-γ^hi NK
IFN-γ^lo NK
IFN-γ^hi MAIT
IFN-γ^lo MAIT
Vα7.2^lo cells
Megakaryocytes
CD8_PAI-1_ADJ
Correlation coefficient (PC) heatmap
Standardized effect size (SES) heatmap
DAD
B.
Correlation coefficient (PC) heatmap
Standardized effect size (SES) heatmap
OP
C.
Correlation coefficient (PC) heatmap
Standardized effect size (SES) heatmap
PC
-1 -0.5 0 0.5 1
SES
-20 -15 -10 -5 0 5 10 15 20

Figure S8

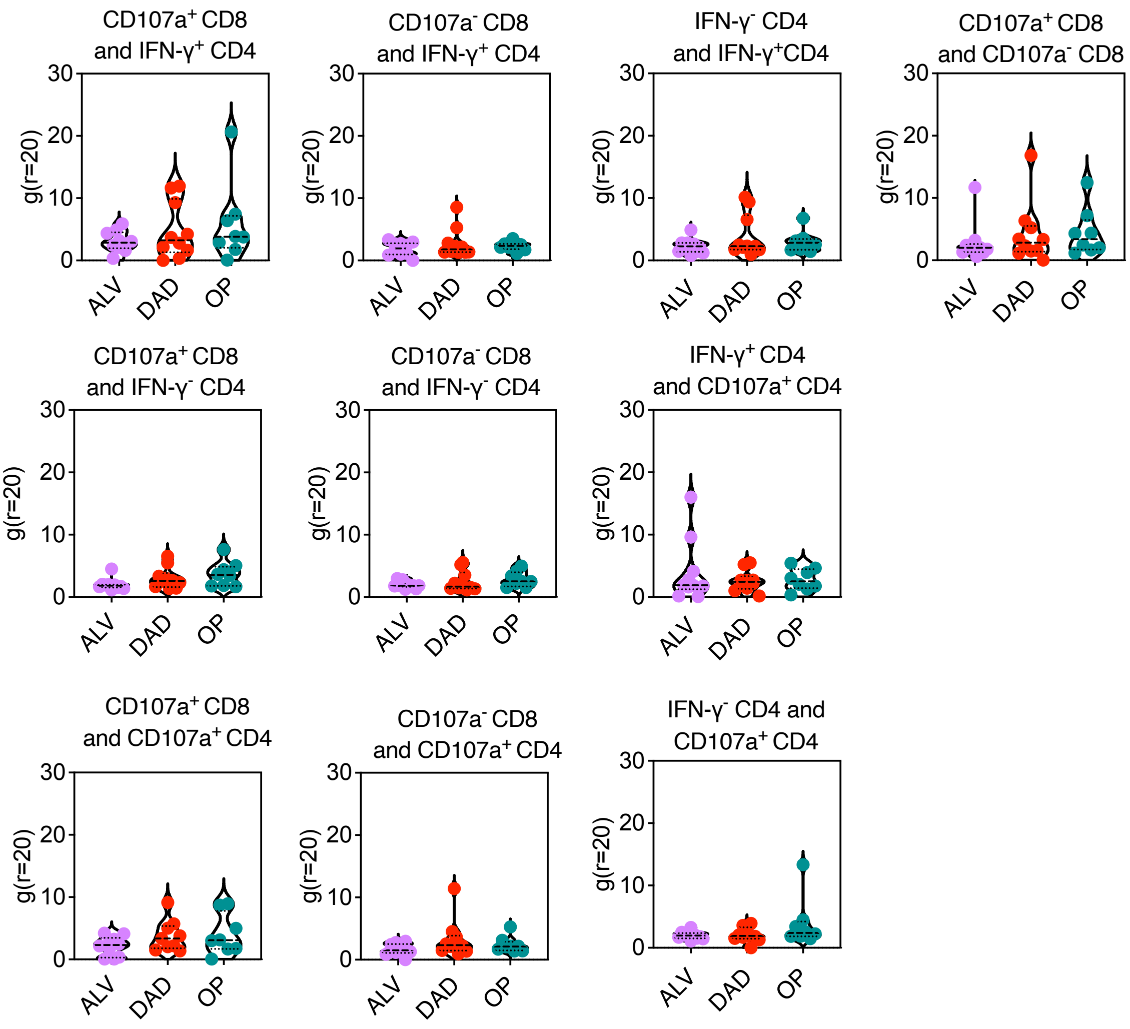
CD107a+ CD8 and IFN-γ+ CD4
CD107a- CD8 and IFN-γ+ CD4
IFN-γ- CD4 and IFN-γ+CD4
CD107a+ CD8 and CD107a- CD8
CD107a+ CD8 and IFN-γ- CD4
CD107a- CD8 and IFN-γ- CD4
IFN-γ+ CD4 and CD107a+ CD4
CD107a+ CD8 and CD107a+ CD4
CD107a- CD8 and CD107a+ CD4
IFN-γ- CD4 and CD107a+ CD4
g(r=20)
0
10
20
30
ALV
DAD
OP

Figure S9

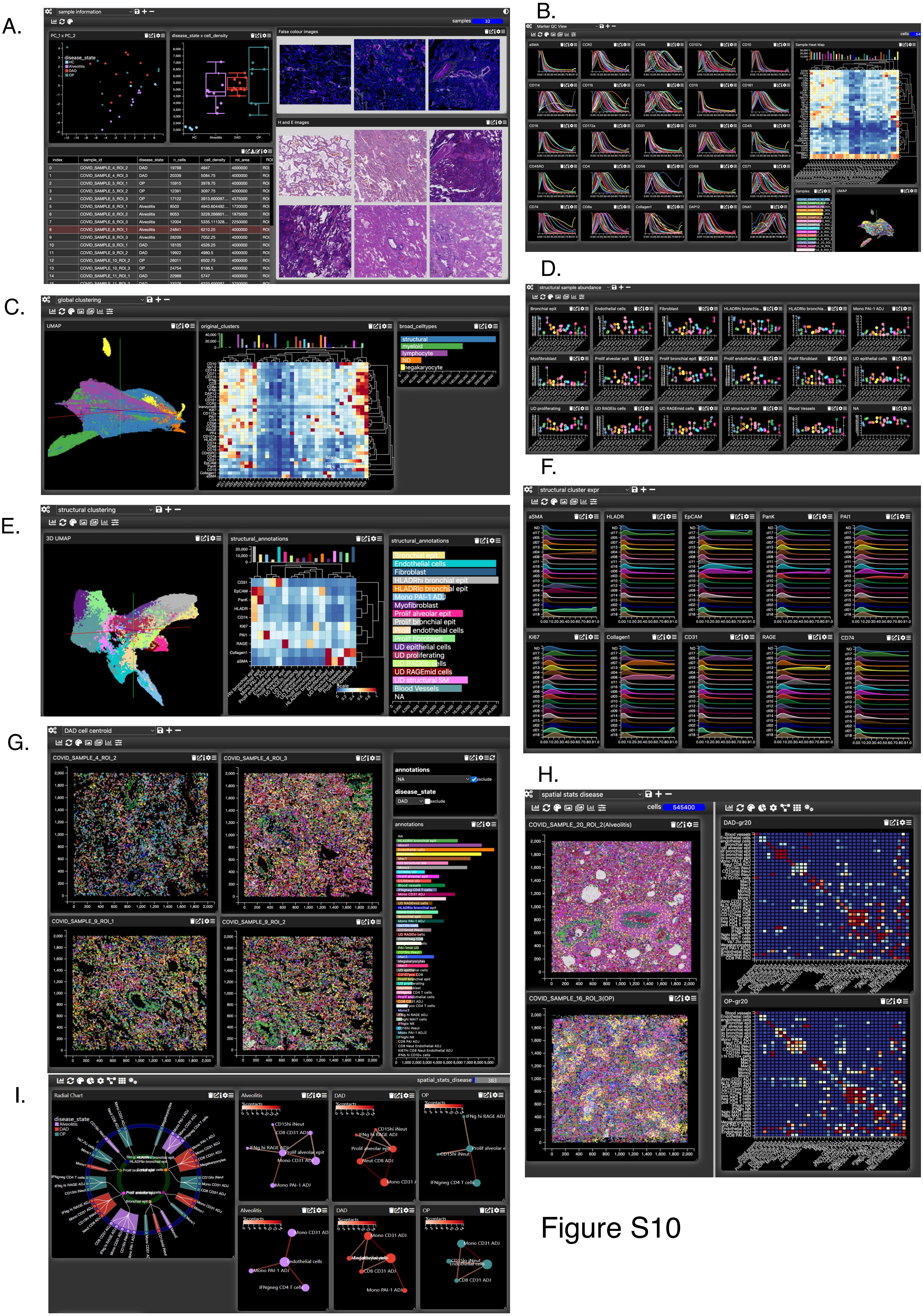
A.
sample information
samples 32
PC_1 x PC_2
disease_state
HC
Alveolitis
DAD
OP
disease_state x cell_density
False colour images
H and E Images
B.
Marker QC View
cells
Sample Heat Map
Samples
UMAP
C.
global clustering
UMAP
original_clusters
broad_celltypes
structural
myeloid
lymphocyte
ND
megakaryocyte
D.
structural sample abundance
E.
structural clustering
3D UMAP
structural_annotations
Bronchial epit
Endothelial cells
Fibroblast
HLADRhi bronchial epit
HLADRlo bronchial epit
Mono PAI-1 ADJ
Myofibroblast
Prolif alveolar epit
Prolif bronchial epit
Prolif endothelial cells
Prolif fibroblast
UD epithelial cells
UD proliferating
UD RAGElo cells
UD RAGEmid cells
UD structural SM
Blood Vessels
NA
F.
structural cluster expr
aSMA
HLADR
EpCAM
PanK
PAI1
Ki67
Collagen1
CD31
RAGE
CD74
G.
DAD cell centroid
COVID_SAMPLE_4_ROI_2
COVID_SAMPLE_4_ROI_3
COVID_SAMPLE_9_ROI_1
COVID_SAMPLE_9_ROI_2
annotations
disease_state
DAD
H.
spatial stats disease
cells 545400
COVID_SAMPLE_20_ROI_2(Alveolitis)
DAD-gr20
COVID_SAMPLE_16_ROI_3(OP)
OP-gr20
I.
spatial_stats_disease
Radial Chart
Alveolitis
DAD
OP

Figure S10

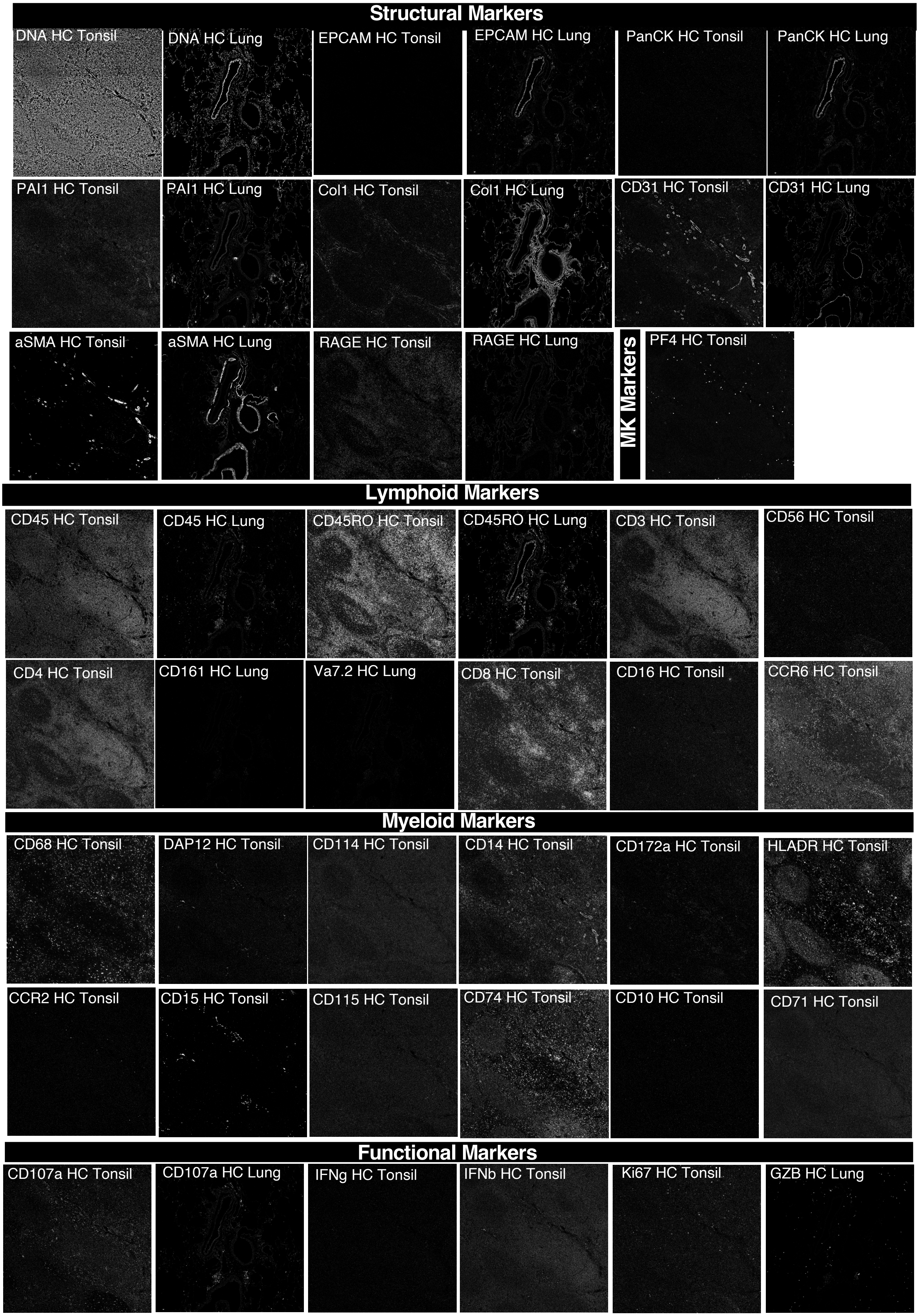
Structural Markers
DNA HC Tonsil
DNA HC Lung
EPCAM HC Tonsil
EPCAM HC Lung
PanCK HC Tonsil
PanCK HC Lung
PAI1 HC Tonsil
PAI1 HC Lung
Col1 HC Tonsil
Col1 HC Lung
CD31 HC Tonsil
CD31 HC Lung
aSMA HC Tonsil
aSMA HC Lung
RAGE HC Tonsil
RAGE HC Lung
MK Markers
PF4 HC Tonsil
Lymphoid Markers
CD45 HC Tonsil
CD45 HC Lung
CD45RO HC Tonsil
CD45RO HC Lung
CD3 HC Tonsil
CD56 HC Tonsil
CD4 HC Tonsil
CD161 HC Lung
Va7.2 HC Lung
CD8 HC Tonsil
CD16 HC Tonsil
CCR6 HC Tonsil
Myeloid Markers
CD68 HC Tonsil
DAP12 HC Tonsil
CD114 HC Tonsil
CD14 HC Tonsil
CD172a HC Tonsil
HLADR HC Tonsil
CCR2 HC Tonsil
CD15 HC Tonsil
CD115 HC Tonsil
CD74 HC Tonsil
CD10 HC Tonsil
CD71 HC Tonsil
Functional Markers
CD107a HC Tonsil
CD107a HC Lung
IFNg HC Tonsil
IFNb HC Tonsil
Ki67 HC Tonsil
GZB HC Lung

Figure S11

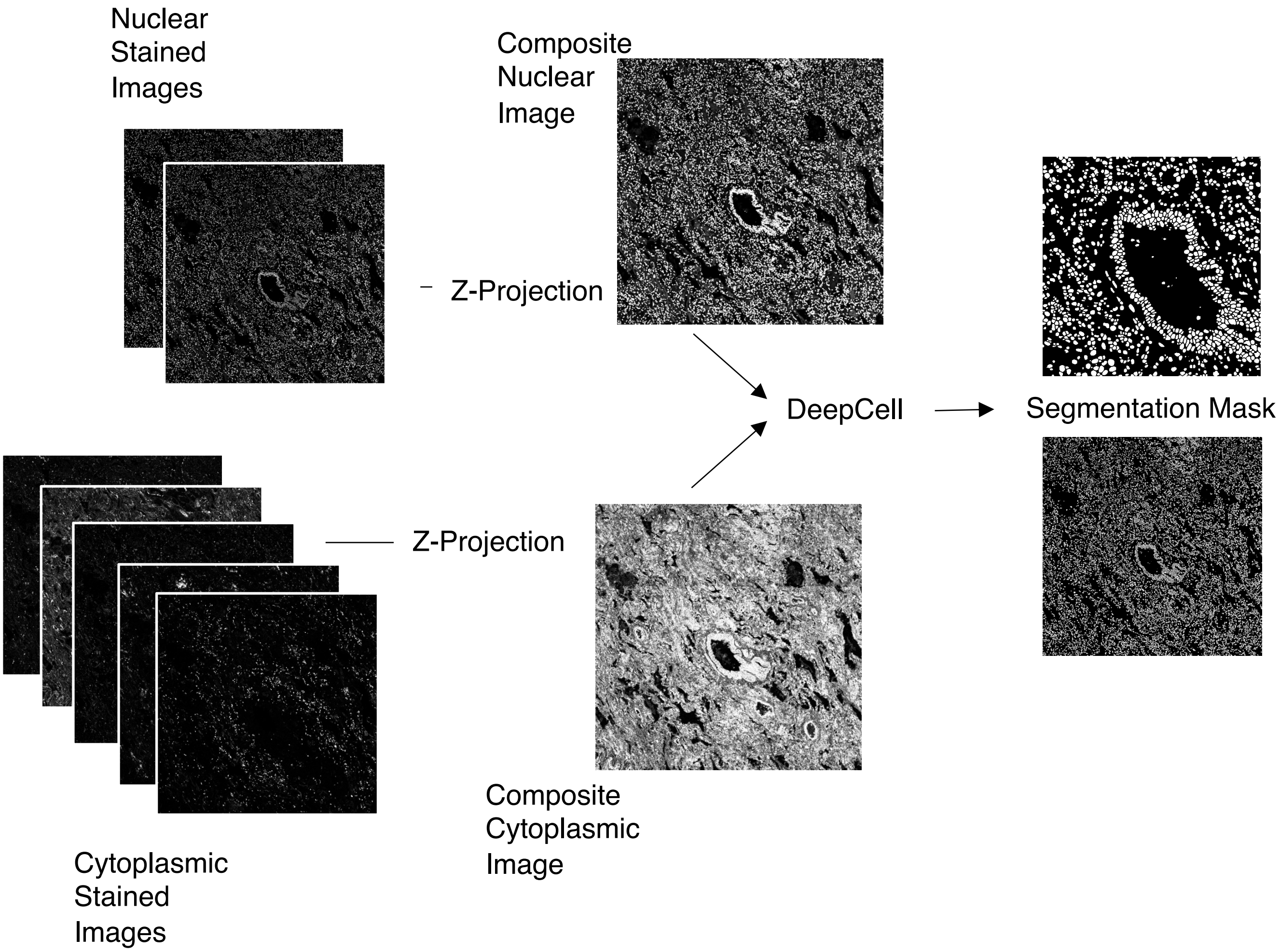
Nuclear Stained Images
Composite Nuclear Image
Z-Projection
DeepCell
Segmentation Mask
Z-Projection
Cytoplasmic Stained Images
Composite Cytoplasmic Image

Figure S12

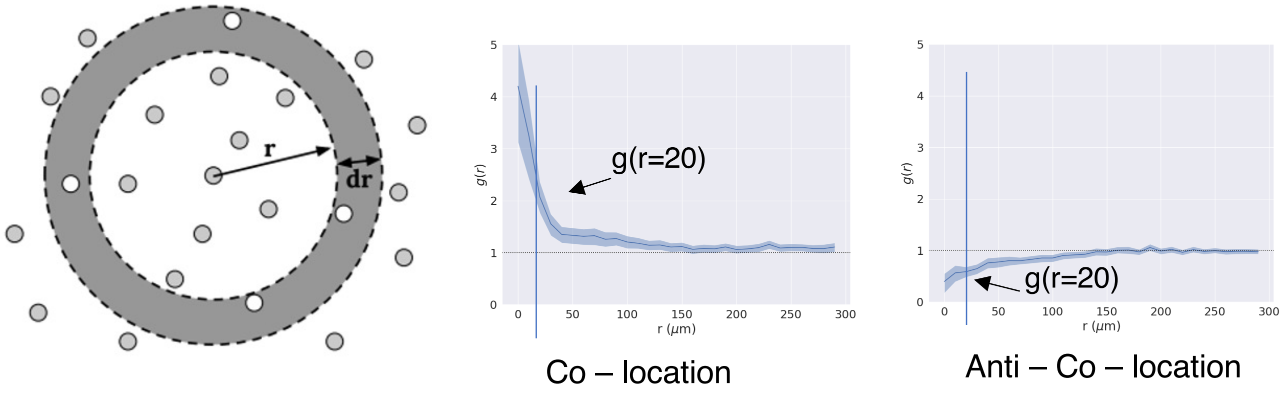


Figure S13

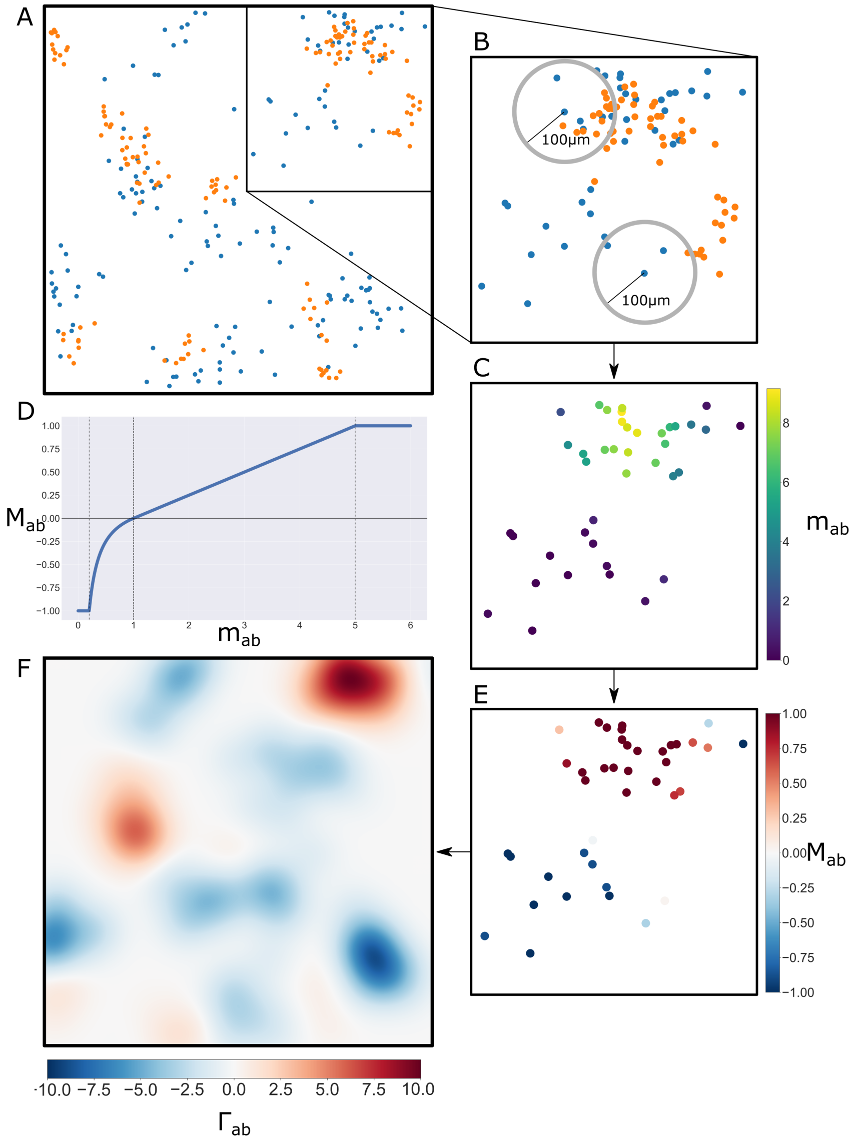


Figure S14

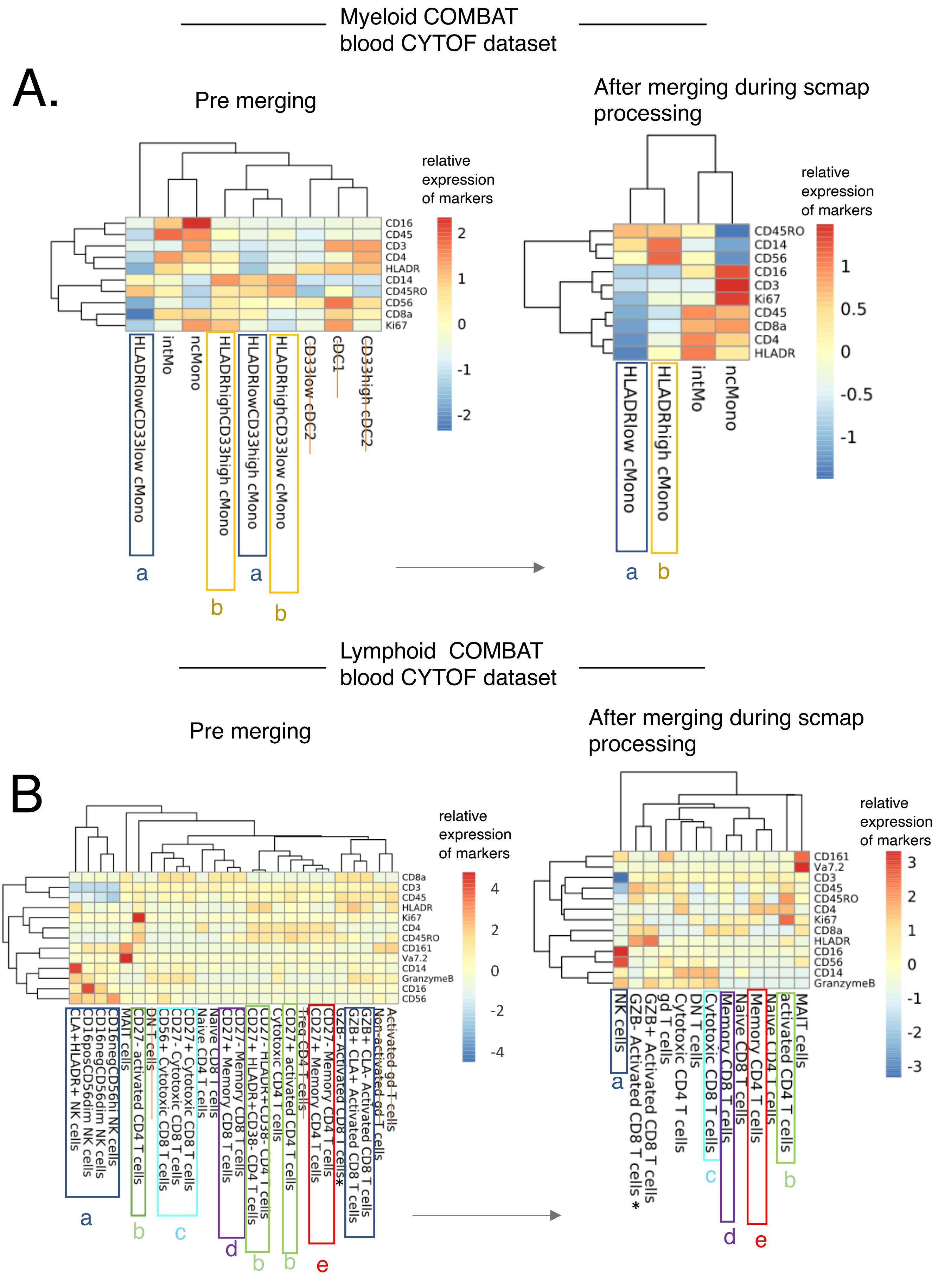
Myeloid COMBAT blood CYTOF dataset
A.
Pre merging
After merging during scmap processing
relative expression of markers
CD16
CD45
CD3
CD4
HLADR
CD14
CD45RO
CD56
CD8a
Ki67
HLADRlowCD33low cMono
intMo
ncMono
HLADRhighCD33high cMono
HLADRlowCD33high cMono
HLADRhighCD33low cMono
CD33low cDC2
cDC1
CD33high cDC2
CD45RO
CD14
CD56
CD16
CD3
Ki67
CD45
CD8a
CD4
HLADR
HLADRlow cMono
HLADRhigh cMono
intMo
ncMono
a
b
Lymphoid COMBAT blood CYTOF dataset
B
Pre merging
After merging during scmap processing
relative expression of markers
CD8a
CD3
CD45
HLADR
Ki67
CD4
CD45RO
CD161
Va7.2
CD14
GranzymeB
CD16
CD56
CLA+HLADR+ NK cells
CD16posCD56dim NK cells
CD16negCD56dim NK cells
CD16negCD56hi NK cells
MAIT cells
CD27- activated CD4 T cells
DN T cells
CD56+ Cytotoxic CD8 T cells
CD27- Cytotoxic CD8 T cells
CD27+ Cytotoxic CD8 T cells
Naive CD4 T cells
Naive CD8 T cells
CD27+ Memory CD8 T cells
CD27- Memory CD8 T cells
CD27+ HLADR+CD38- CD4 T cells
CD27- HLADR+CD38- CD4 T cells
Cytotoxic CD4 T cells
CD27+ activated CD4 T cells
Treg CD4 T cells
CD27+ Memory CD4 T cells
CD27- Memory CD4 T cells
GZB- Activated CD8 T cells*
GZB+ CLA+ Activated CD8 T cells
GZB+ CLA- Activated CD8 T cells
Non-activated gd T cells
Activated gd T cells
CD161
Va7.2
CD3
CD45
CD45RO
CD4
Ki67
CD8a
HLADR
CD16
CD56
CD14
GranzymeB
NK cells
GZB- Activated CD8 T cells *
GZB+ Activated CD8 T cells
gd T cells
Cytotoxic CD4 T cells
DN T cells
Cytotoxic CD8 T cells
Memory CD8 T cells
Naive CD8 T cells
Memory CD4 T cells
Naive CD4 T cells
activated CD4 T cells
MAIT cells
a
b
c
d
e

Figure S15
