## Supplementary material for "Unbiased single cell spatial analysis localises inflammatory clusters of immature neutrophils-CD8 T cells to alveolar progenitor cells in fatal COVID-19 lungs": Suppl Table 3

### Slide 1
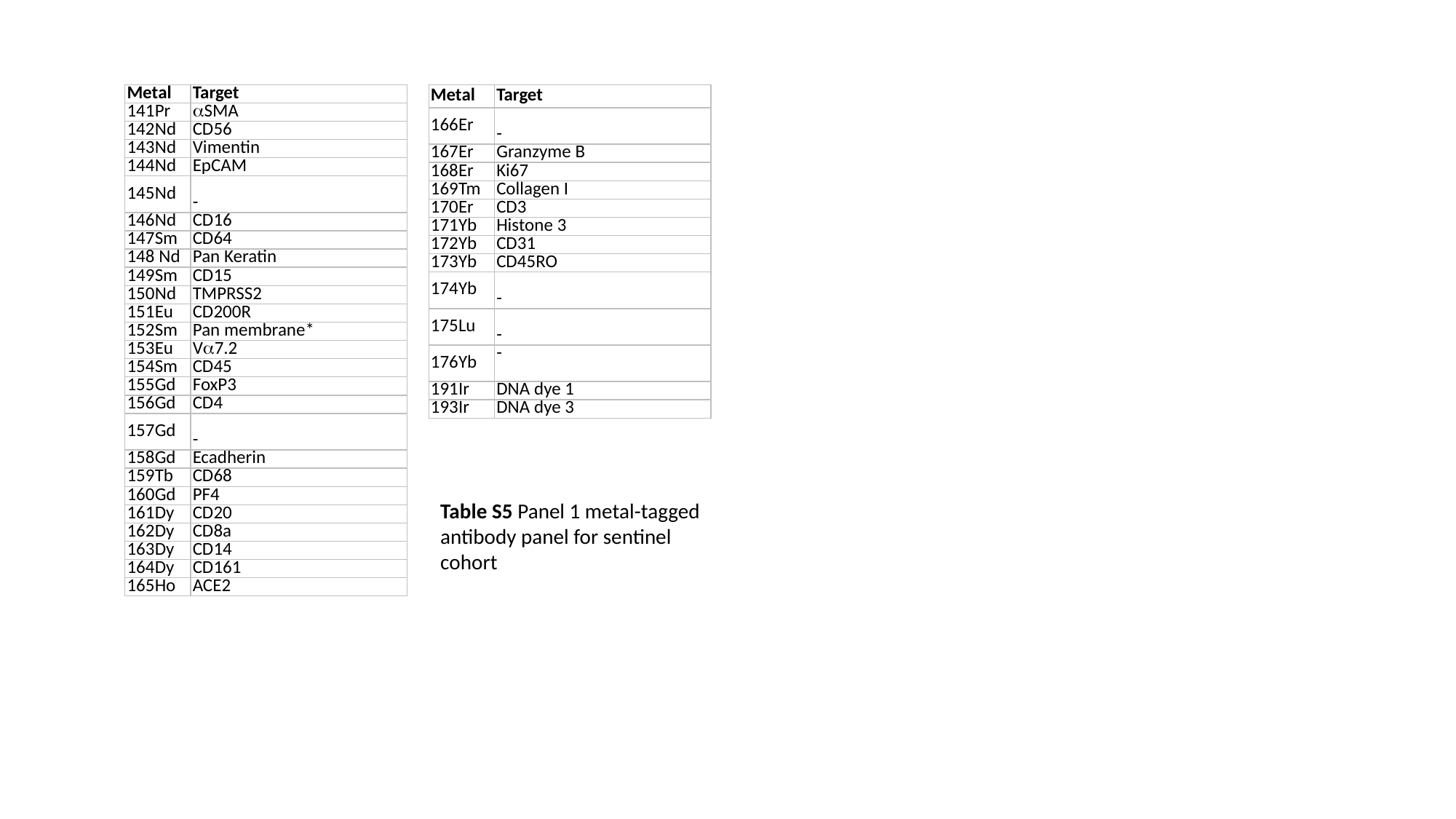

| Metal | Target |
| --- | --- |
| 141Pr | αSMA |
| 142Nd | CD56 |
| 143Nd | Vimentin |
| 144Nd | EpCAM |
| 145Nd | - |
| 146Nd | CD16 |
| 147Sm | CD64 |
| 148 Nd | Pan Keratin |
| 149Sm | CD15 |
| 150Nd | TMPRSS2 |
| 151Eu | CD200R |
| 152Sm | Pan membrane\* |
| 153Eu | Vα7.2 |
| 154Sm | CD45 |
| 155Gd | FoxP3 |
| 156Gd | CD4 |
| 157Gd | - |
| 158Gd | Ecadherin |
| 159Tb | CD68 |
| 160Gd | PF4 |
| 161Dy | CD20 |
| 162Dy | CD8a |
| 163Dy | CD14 |
| 164Dy | CD161 |
| 165Ho | ACE2 |
| Metal | Target |
| --- | --- |
| 166Er | - |
| 167Er | Granzyme B |
| 168Er | Ki67 |
| 169Tm | Collagen I |
| 170Er | CD3 |
| 171Yb | Histone 3 |
| 172Yb | CD31 |
| 173Yb | CD45RO |
| 174Yb | - |
| 175Lu | - |
| 176Yb | - |
| 191Ir | DNA dye 1 |
| 193Ir | DNA dye 3 |
Table S5 Panel 1 metal-tagged
antibody panel for sentinel
cohort
