## Supplementary material for "Unbiased single cell spatial analysis localises inflammatory clusters of immature neutrophils-CD8 T cells to alveolar progenitor cells in fatal COVID-19 lungs": Suppl Table 5

### Slide 1
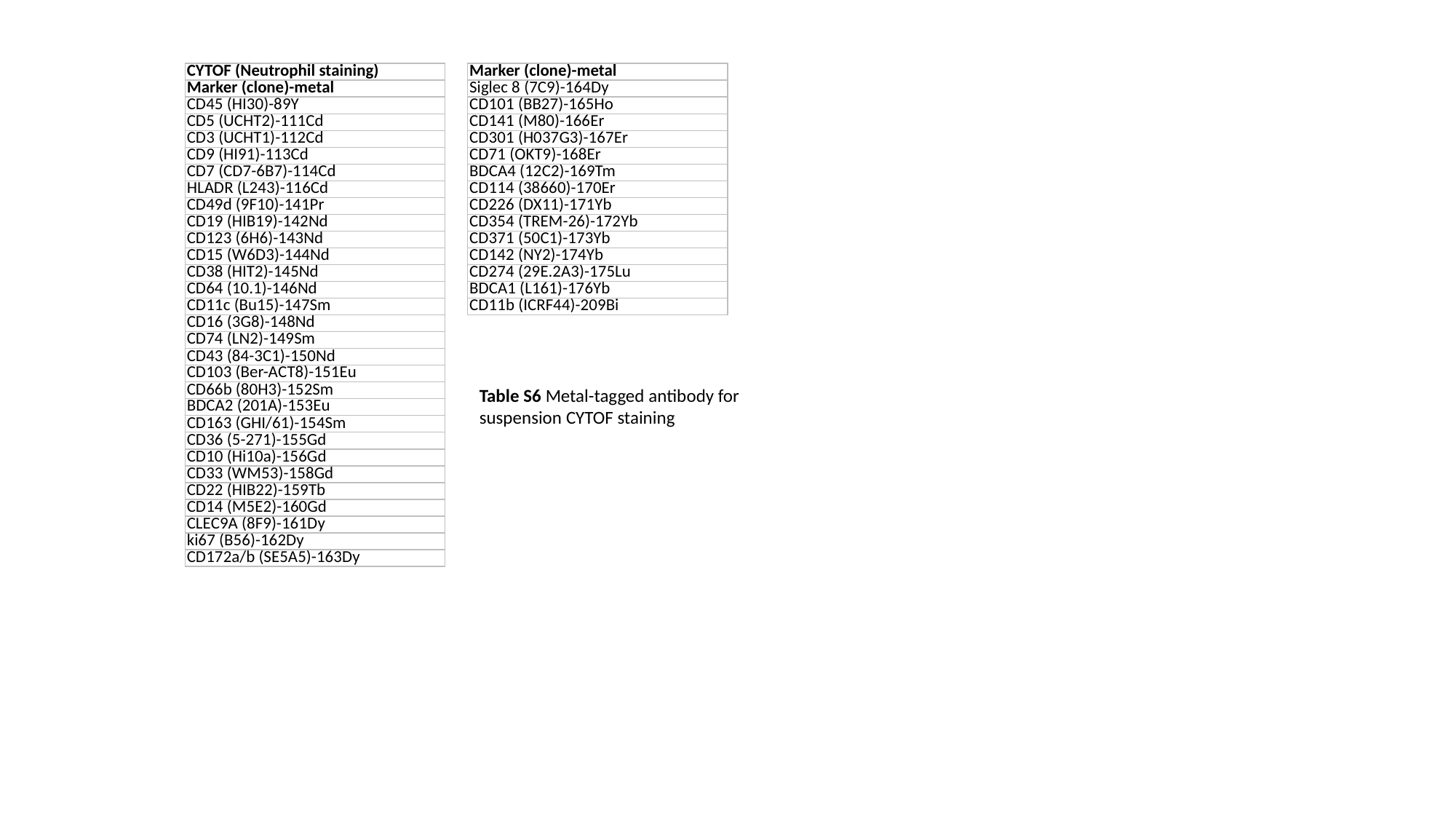

| CYTOF (Neutrophil staining) |
| --- |
| Marker (clone)-metal |
| CD45 (HI30)-89Y |
| CD5 (UCHT2)-111Cd |
| CD3 (UCHT1)-112Cd |
| CD9 (HI91)-113Cd |
| CD7 (CD7-6B7)-114Cd |
| HLADR (L243)-116Cd |
| CD49d (9F10)-141Pr |
| CD19 (HIB19)-142Nd |
| CD123 (6H6)-143Nd |
| CD15 (W6D3)-144Nd |
| CD38 (HIT2)-145Nd |
| CD64 (10.1)-146Nd |
| CD11c (Bu15)-147Sm |
| CD16 (3G8)-148Nd |
| CD74 (LN2)-149Sm |
| CD43 (84-3C1)-150Nd |
| CD103 (Ber-ACT8)-151Eu |
| CD66b (80H3)-152Sm |
| BDCA2 (201A)-153Eu |
| CD163 (GHI/61)-154Sm |
| CD36 (5-271)-155Gd |
| CD10 (Hi10a)-156Gd |
| CD33 (WM53)-158Gd |
| CD22 (HIB22)-159Tb |
| CD14 (M5E2)-160Gd |
| CLEC9A (8F9)-161Dy |
| ki67 (B56)-162Dy |
| CD172a/b (SE5A5)-163Dy |
| Marker (clone)-metal |
| --- |
| Siglec 8 (7C9)-164Dy |
| CD101 (BB27)-165Ho |
| CD141 (M80)-166Er |
| CD301 (H037G3)-167Er |
| CD71 (OKT9)-168Er |
| BDCA4 (12C2)-169Tm |
| CD114 (38660)-170Er |
| CD226 (DX11)-171Yb |
| CD354 (TREM-26)-172Yb |
| CD371 (50C1)-173Yb |
| CD142 (NY2)-174Yb |
| CD274 (29E.2A3)-175Lu |
| BDCA1 (L161)-176Yb |
| CD11b (ICRF44)-209Bi |
Table S6 Metal-tagged antibody for
suspension CYTOF staining
