## Supplementary material for "Unbiased single cell spatial analysis localises inflammatory clusters of immature neutrophils-CD8 T cells to alveolar progenitor cells in fatal COVID-19 lungs": Suppl Table 4

### Slide 1
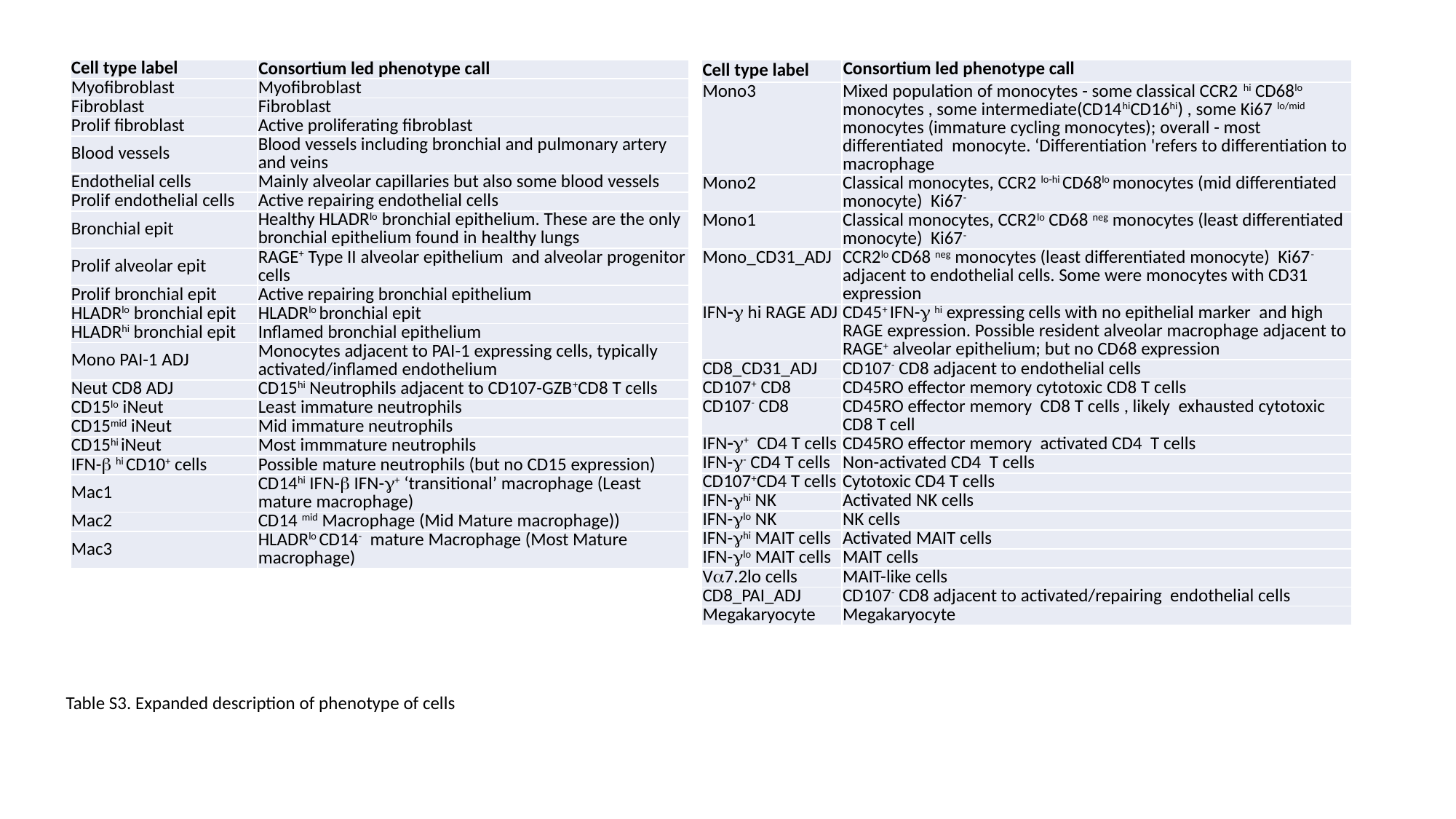

| Cell type label | Consortium led phenotype call |
| --- | --- |
| Myofibroblast | Myofibroblast |
| Fibroblast | Fibroblast |
| Prolif fibroblast | Active proliferating fibroblast |
| Blood vessels | Blood vessels including bronchial and pulmonary artery and veins |
| Endothelial cells | Mainly alveolar capillaries but also some blood vessels |
| Prolif endothelial cells | Active repairing endothelial cells |
| Bronchial epit | Healthy HLADR^lo^ bronchial epithelium. These are the only bronchial epithelium found in healthy lungs |
| Prolif alveolar epit | RAGE^+^ Type II alveolar epithelium and alveolar progenitor cells |
| Prolif bronchial epit | Active repairing bronchial epithelium |
| HLADR^lo^ bronchial epit | HLADR^lo^ bronchial epit |
| HLADR^hi^ bronchial epit | Inflamed bronchial epithelium |
| Mono PAI-1 ADJ | Monocytes adjacent to PAI-1 expressing cells, typically activated/inflamed endothelium |
| Neut CD8 ADJ | CD15^hi^ Neutrophils adjacent to CD107-GZB^+^CD8 T cells |
| CD15^lo^ iNeut | Least immature neutrophils |
| CD15^mid^ iNeut | Mid immature neutrophils |
| CD15^hi^ iNeut | Most immmature neutrophils |
| IFN-β ^hi^ CD10^+^ cells | Possible mature neutrophils (but no CD15 expression) |
| Mac1 | CD14^hi^ IFN-β IFN-γ^+^ ‘transitional’ macrophage (Least mature macrophage) |
| Mac2 | CD14 ^mid^ Macrophage (Mid Mature macrophage)) |
| Mac3 | HLADR^lo^ CD14^-^ mature Macrophage (Most Mature macrophage) |
| Cell type label | Consortium led phenotype call |
| --- | --- |
| Mono3 | Mixed population of monocytes - some classical CCR2 ^hi^ CD68^lo^ monocytes , some intermediate(CD14^hi^CD16^hi^) , some Ki67 ^lo/mid^ monocytes (immature cycling monocytes); overall - most differentiated monocyte. ‘Differentiation 'refers to differentiation to macrophage |
| Mono2 | Classical monocytes, CCR2 ^lo-hi^ CD68^lo^ monocytes (mid differentiated monocyte) Ki67^-^ |
| Mono1 | Classical monocytes, CCR2^lo^ CD68 ^neg^ monocytes (least differentiated monocyte) Ki67^-^ |
| Mono\_CD31\_ADJ | CCR2^lo^ CD68 ^neg^ monocytes (least differentiated monocyte) Ki67^-^ adjacent to endothelial cells. Some were monocytes with CD31 expression |
| IFN−γ hi RAGE ADJ | CD45^+^ IFN-γ ^hi^ expressing cells with no epithelial marker and high RAGE expression. Possible resident alveolar macrophage adjacent to RAGE^+^ alveolar epithelium; but no CD68 expression |
| CD8\_CD31\_ADJ | CD107^-^ CD8 adjacent to endothelial cells |
| CD107^+^ CD8 | CD45RO effector memory cytotoxic CD8 T cells |
| CD107^-^ CD8 | CD45RO effector memory CD8 T cells , likely exhausted cytotoxic CD8 T cell |
| IFN−γ^+^ CD4 T cells | CD45RO effector memory activated CD4 T cells |
| IFN-γ^-^ CD4 T cells | Non-activated CD4 T cells |
| CD107^+^CD4 T cells | Cytotoxic CD4 T cells |
| IFN-γ^hi^ NK | Activated NK cells |
| IFN-γ^lo^ NK | NK cells |
| IFN-γ^hi^ MAIT cells | Activated MAIT cells |
| IFN-γ^lo^ MAIT cells | MAIT cells |
| Vα7.2lo cells | MAIT-like cells |
| CD8\_PAI\_ADJ | CD107^-^ CD8 adjacent to activated/repairing endothelial cells |
| Megakaryocyte | Megakaryocyte |
Table S3. Expanded description of phenotype of cells
